# Level of clinical competence in junior medical residents and its correlation with antibiotic prescription errors: a cross-sectional study

**DOI:** 10.1101/2020.04.28.20083584

**Authors:** Martínez D. Joshua, Sierra-Martínez Octavio, Galindo-Fraga Arturo, Trejo Mejía Juan Andrés, Sánchez-Mendiola Melchor, Ochoa-Hein Eric, Vázquez-Rivera Mirella, Gutiérrez-Cirlos Carlos, Naveja Jesús, Martínez-González Adrián

## Abstract

**Background:** A large portion of prescribing errors can be attributed to medication knowledge deficiency. They are preventable and most often occur in the stage of ordering. Antimicrobials are the drug class most commonly related to prescribing errors.

**Objectives:** The study main objective was to describe the relationship between clinical competence and antibiotic prescription errors. Secondary objectives were to measure clinical competence of junior medical residents with an Objective Structured Clinical Examination (OSCE), to describe the frequency and severity of antibiotic prescription errors and to find items and attributes of clinical competence that are correlated with the antibiotic prescription error ratio.

**Methods:** A cross-sectional study was designed to assess the clinical competence of junior medical residents, from National Institute of Pediatrics and “Manuel Gea Gonzalez” General Hospital in Mexico City, through an infectious disease OSCE and measure the frequency and severity of antibiotic prescription errors. Statistical analysis included generalizability theory and internal consistency Cronbach’s alpha, a partial correlation controlling sex and time of degree, simple linear regression and item’s exploratory factorial analysis.

**Results:** The mean OSCE score was 0.692 ± 0.073. The inter-item Cronbach’s alpha was 0.927 and inter-station Cronbach’s alpha was 0.774. The G coefficient in generalizability theory analysis was 0.84. The antibiotic prescription error ratio was 45.1% ± 7%. The severity of antibiotic prescription errors was: category C (errors that do not cause patient harm) = 56 cases, 15.5%; category D (monitoring required to confirm that errors resulted in no harm to the patient or intervention required to preclude harm) = 51 cases, 14.1%; category E (errors that may contribute to or result in temporary harm to the patient and require intervention) = 235, 65.2%; category F (errors that may contribute to or resulted in temporary harm to the patient and require initial or prolonged hospitalization) = 18 cases, 5%. The correlation between clinical competence and antibiotic prescription errors was established with Pearson correlation (r=-0.33, p<0.05, CI95% -0.57 to -0.07), and partial correlation controlling effect of gender and time since graduation (r=-0.39, p<0.01, CI95% -0.625 to -0.118).

**Conclusions:** We found a negative correlation between clinical competence and antibiotic prescription error ratio in graduated physicians who have been accepted in a medical specialty. The therapeutic plan, which is a component of clinical competence score, and the prescription skills had a negative correlation with antibiotic prescription errors. The most frequent mistakes in antibiotic prescriptions errors would need a second intervention.

## Background

Medical prescribing errors may be defined based on the causes, processes, and outcomes. One definition includes all of these: “an act of omission or commission in planning or execution that contributes or could contribute to an unintended result” (1). In addition, there are many types of errors beyond those regarding medication, such as surgical mistakes or skill deficiencies, as well as misdiagnoses (2). We focused on prescription errors, one subtype of medication errors.

The most commonly incorrectly prescribed drug class are antimicrobials (3–5). The majority (80%) of antibiotic prescribing takes place in the community and injudicious use of antibiotics is a major factor facilitating the emergence of resistance worldwide (6). Furthermore, antimicrobial resistance is related to the amount of antibiotic consumed and the class of antibiotics (7).

The early empirical antimicrobial treatment is associated with a significant reduction in all-cause mortality (8). Inappropriate initial antimicrobial therapy for septic shock occurs in about 20% of patients and is associated with a fivefold reduction in survival (9).

A large portion of prescribing errors can be attributed to medication knowledge deficiency (3). Determining the role of lack of medication knowledge as a proximal cause of errors is important in designing error prevention strategies (4). Despite a 50% decrease in preventable adverse drug events with computerized provider order entry (10), this system didn’t reduce the prescription errors and may only be determinant in pharmacy transcription or validation errors, as well as nursing transcription and dispensation errors (11).

Competence in medicine is defined by Epstein as “the habitual and judicious use of communication, knowledge, technical skills, clinical reasoning, emotions, values and reflection in daily practice for the benefit of the individuals and communities being served” (12) and includes a set of attributes like clinical skills, knowledge, interpersonal skills, problem solving, clinical judgment and technical skills (13). Competence is contextual, reflecting the relationship between abilities and the tasks required to perform in a particular situation in the real world.

Assessment of clinical competence can be done with different methods. One of them is the Objective Structured Clinical Examination (OSCE) developed by Harden (14, 15), considered the instrument of choice to assess competency in health care professionals (16–18). The reliability and validity of an OSCE increases with the number of stations, although other factors might be involved (19).

We designed and applied an OSCE to assess clinical competence of first-year medical residents in ambulatory patients with infectious diseases.

## Objective

To describe the relationship between clinical competence and antibiotic prescription errors in first-year medical residents in the settings of an OSCE.

We also measured the frequency and severity of antibiotics prescription errors. Therefore, we identified items and attributes of clinical competence that were associated with antibiotics prescription error ratio.

## Method

### Study design

A cross-sectional study using an OSCE was designed and applied to first-year medical residents at three medical institutions in Mexico City in February 2019.

The participants were assessed in nine OSCE stations: Pulmonary tuberculosis, Acute Pyelonephritis, Latent Syphilis, Community acquired pneumonia, Acute pharyngitis, Acute gastroenteritis, Gonorrheal urethritis, Cellulitis and Acute Cystitis. Stations were dynamic, had one rater, one standardized patient with an infectious disease clinical case. The complexity of the clinical case was designed targeted to the knowledge level of a general physician. The infectious disease cases were selected according to their outpatient prevalence. Each case and its related treatment were approved by independent consensus of two Infectious Disease physicians, members of the Infectiology and clinical microbiology Mexican association in active clinical practice. We took into consideration both the local antibiotic resistance patterns as well as the suggested empirical treatments as per national and international guidelines (20–51).

Furthermore, the solutions to the clinical cases regarding the diagnosis were consistent with a medical diagnostic decision support system (DXplain™) (52,53); the right answers regarding the treatment were determined by the standards of care of the most likely diagnosis in each case. All stations were tested and improved with family medicine residents.

### Settings

The medical institutions selected were “Manuel Gea Gonzalez” General Hospital, “Salvador Zubiran” National Institute of Medical Sciences and Nutrition and the National Institute of Pediatrics, all of them in Mexico City during February 2019, before the beginning of the specialty courses.

### Participants

First year medical residents in the selected institutions and belonging to a direct-entry medical specialty (General Surgery, Gynecology, Internal Medicine or Pediatrics) were invited to participate. Residents that had more than 12 hours of continuous work were excluded from the study.

### Raters

A total of 18 raters from Faculty of medicine, National Autonomous University of Mexico (UNAM) took part in the assess process. All raters were physicians and had taken an OSCE workshop course of 15 hours where they developed one OSCE station. The raters reviewed the stations and the guidelines for the examiner before the test. In addition, they were updated with the antibiotic treatments of stations. Each rater used an op-scan sheet to score each student with the rating scale.

Raters had experience with OSCE (14 raters with 10 years of experience and 4 raters with 3 years of experience). What’s more, they participated in 3–4 OSCEs each year.

### Standardized patients

A total of 18 standardized patients participated in the examination. All standardized patients taken an OSCE workshop course of 4 hours and an acting course of 4 hours. They had a similar age according to clinical cases. Moreover, they were provided with a dialogue guideline that includes patient personality.

Standardized patients had experience with OSCE (6 patients with 10 years of experience and 12 patients with 3 years of experience). In addition, they participated in 3–4 OSCEs each year.

### Variables

The independent variables were clinical competence, age, sex, time elapsed since graduation of medical school, global assessment of knowledge and prescription skills. The dependent variables were antibiotic prescription error ratio (proportion of antibiotics errors) and the severity of prescription errors.

### Instrument

We designed an OSCE that included the guidelines for the examiner, reported by Martinez-Gonzalez et al. with Cronbach’s alpha = 0.94 (54). The guidelines and rating scales (54) for each clinical competence component included anamnesis, physical examination, laboratory and imaging tests, diagnosis, therapeutic plan, communication, and patient’s assessment.

Patient’s assessment is important to measure the interpersonal skills. Answer the question “How the physician makes feel to patient?” and range since distrust, apathy, coldness, mistreatment to trust, empathy, attention and kindness.

The prescription and global assessment of knowledge and skills items were measured with rating scales at every station. The prescription item allowed to the rater assess the quality of medical prescription document, written by the junior medical resident, and range since wrong drug to right in all “5 rights”.

Antibiotic prescription errors were measured with the *“five rights”* (right patient, right drug, right dose, right route, and right time) using the information that the resident wrote (55). The severity of prescription errors was measured with the National Coordinating Council for Medication Errors Reporting and Prevention (NCC MERP) index, which has acceptable validity and reliability (56).

### Bias management

Most observational studies of prescription errors have found no difference when adjusting the results by the academic level of the physicians, environmental factors at the moment of prescription, or the difficulty and type of clinical cases. This OSCE was designed to simultaneously assess the clinical competence and prescription errors in junior medical residents, adjusting by time elapsed since graduation. Furthermore, we considered the local antibiotic resistance of etiologic agents, avoided including fatigued physicians and restricted the access to medical databases during the assessment process.

Following the results of other OSCEs (17,57) that detected sources of errors through G theory, we designed this OSCE to achieve minimal sources of errors by including trained examiners and standardized patients. The assessment was applied in 2 different days depending on the institution in the same schedule. Each clinical case and its treatment were approved by consensus of two independent Infectious Disease physicians.

### Sample size

The sample size consisted of 51 medical residents. A sample size of 49 medical residents is sufficient to show correlation between the main variables with r= 0.39, α= 0.05 and 80% of statistical power (58).

### Statistical analysis

The guidelines for examiners had a rating scale for each clinical competence component: 0.25, insufficient; 0.5, adequate; 0.75, good and 1, excellent. The weighting of the clinical competence score was anamnesis, 30%; physical examination, 12%; laboratory and imaging testing, 16%; diagnosis, 12%; therapeutic plan, 12%; communication, 12% and patient, 6%. Global assessment of knowledge and prescription skills didn’t contribute to the clinical competence score.

The internal consistency, both inter-station and inter-item, was measured with Cronbach’s alpha. The generalizability was measured with the G coefficient. We used the estimated variance components for each of the following facts: stations, day of the assessment, medical resident and all their interactions (17).

The clinical competence and antibiotic prescription error proportion were assessed with Anderson-Darling, D’Agostino & Pearson, and Shapiro-Wilk normality test (52,53). To describe the relation between clinical competence and antibiotic prescription error ratio we used the Pearson correlation test and simple linear regression. The Spearman’s correction for attenuation was applied between clinical competence and antibiotic prescription error ratio(59). An exploratory factorial analysis (EFA) was applied to describe the factors of clinical competence that were correlated with antibiotic prescription errors, using maximum likelihood extraction and equamax rotation method.

Data analysis was conducted using IBM SPSS 25 and JMP 11 SAS software.

### Ethical aspects

This study was approved by research and ethics committees of three institutions: “Manuel Gea Gonzalez” General Hospital (approval no. 39-26-2018), “Salvador Zubirán” National Institute of Medical Sciences and Nutrition (approval no. 2863) and the National Autonomous University of Mexico (UNAM) Faculty of Medicine, (approval no. 021/PECEM/2018). The identity of the residents was anonymized by masking, pseudo-anonymization, and aggregation.

## Results

### Participants

The OSCE was applied to 51 medical residents in February 2019. The characteristics of junior residents are showed in Table 1.

**Table 1.** Participating junior residents characteristics (n=51)

| Characteristic | Value (%), Mean ± SD or Median (Q<sub>1</sub>-Q<sub>3</sub>) |
| --- | --- |
| Men | 20 (39.2%) |
| Female | 31 (60.8%) |
| Nacional Institute of Pediatrics | 44 (86.3%) |
| "Manuel Gea Gonzalez" General Hospital | 7 (13.7%) |
| Pediatrics | 41 (80.4%) |
| Medical genetics | 3 (5.9%) |
| Internal medicine | 2 (3.9%) |
| Traumatology and orthopedics | 5 (9.8%) |
| ENARM* score | 78.082 ± 3.03 |
| ENARM* highest score | 84 |
| ENARM* lowest score | 71.7 |
| ENARM* 1 time | 31 (60.8%) |
| ENARM* 2 times | 12 (23.5%) |
| ENARM* 3 times | 8 (15.7%) |
| Months after graduation from medical school | 7 (5-18) |
| *National Test for Aspirants to Medical Residency (ENARM) |  |

### OSCE results

The medical residents mean OSCE score was 0.692 ± 0.073 SD (Figure 1). This score is an average proportion of correct answers in the test. Clinical competence component scores are: Anamnesis 0.682 ± 0.091, Physical examination 0.686 ± 0.093, Lab and imaging test 0.693 ± 0.099, Diagnosis 0.686 ± 0.079, Therapeutic plan 0.604 ± 0.097, Communication 0.778 ± 0.099 and Patient 0.781 ± 0.098. In addition, we showed items additional measured items: Prescription 0.496 ± 0.080 and Global assessment of knowledge and skills 0.644 ± 0.082. The OSCE score had a normal distribution. The inter-item Cronbach’s alpha was 0.927 and inter station Cronbach’s alpha was 0.774. The G coefficient from the generalizability analysis was 0.84.

**Figure 1.**
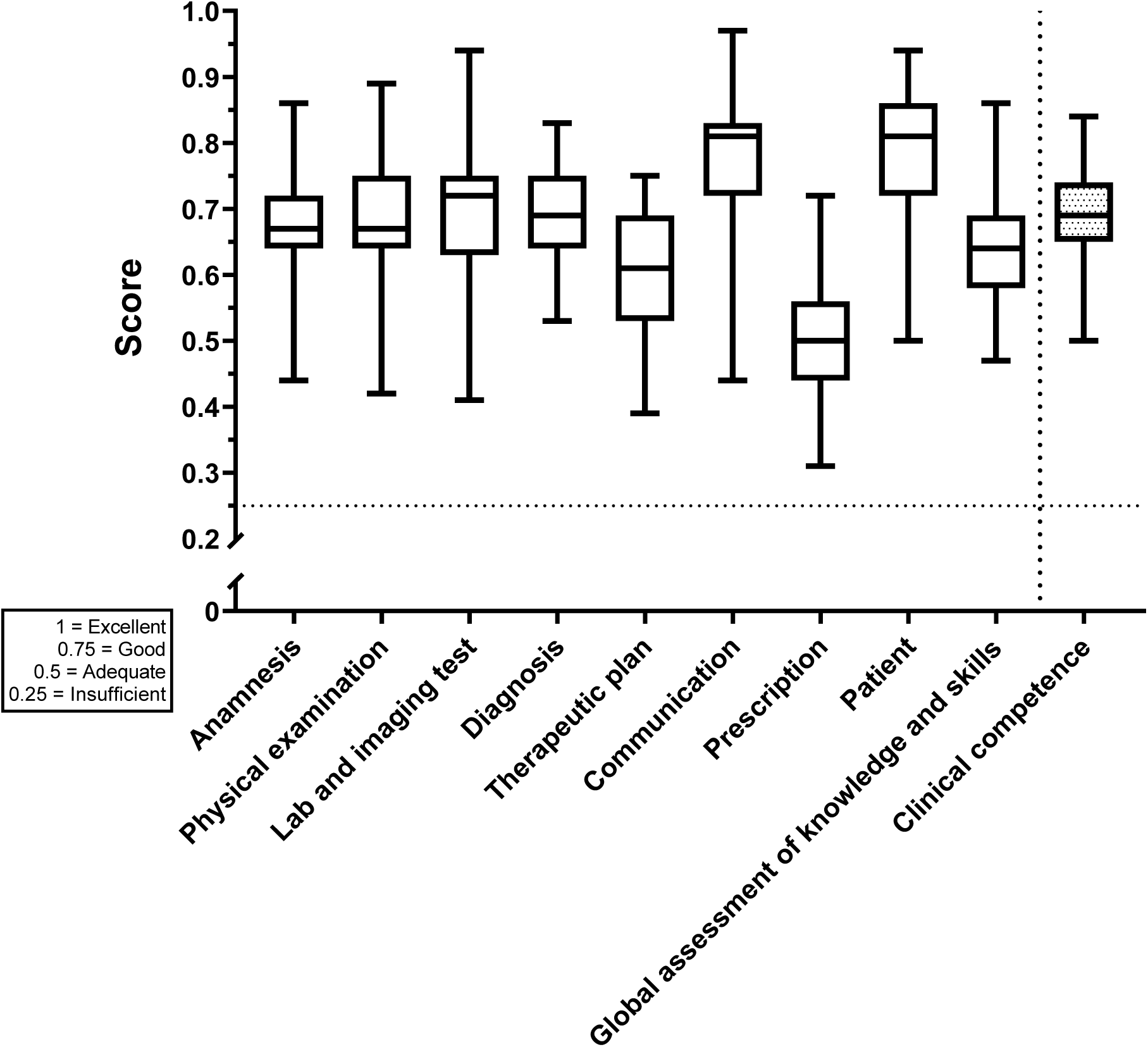
Clinical competence and its components in junior residents (n=51) in an infectious disease OSCE. The score scale is 1 = Excellent, 0.75 = Good, 0.5 = Adequate and 0.25 = Insufficient.

The antibiotic prescription error ratio was 0.451 ± 0.07 SD. Higher rates of prescription errors were observed regarding doses, administration route and time. The antibiotic prescription error ratio for each station is depicted in Figure 2.

**Figure 2.**
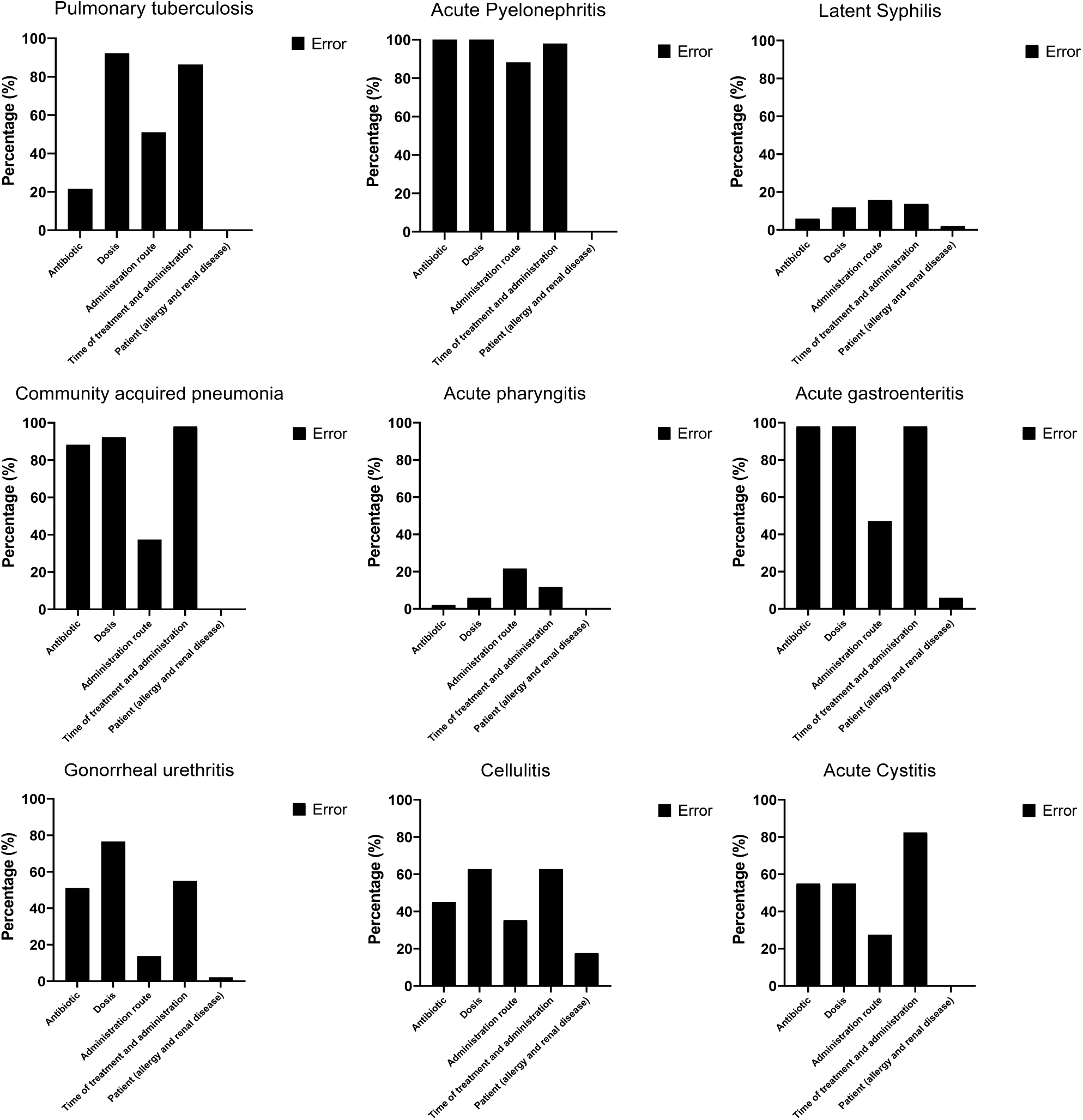
Antibiotic prescription errors ratio using “the 5 rights” (antibiotic, dose, administration route, time of treatment and administration and patient) in each OSCE station.

The severity of antibiotic prescription errors was: category C (errors that do not cause patient harm) = 56 cases, 15.5%; category D (monitoring required to confirm that errors resulted in no harm to the patient or intervention required to preclude harm) = 51 cases, 14.1%; category E (errors that may contribute to or result in temporary harm to the patient and require intervention) = 235, 65.2%; category F (errors that may contribute to or resulted in temporary harm to the patient and require initial or prolonged hospitalization) = 18 cases, 5%. Categories A, B, G, H and I from the NCC MERP index (56) were not considered in this study. The severity of prescription errors at each station is shown in Figure 3.

**Figure 3.**
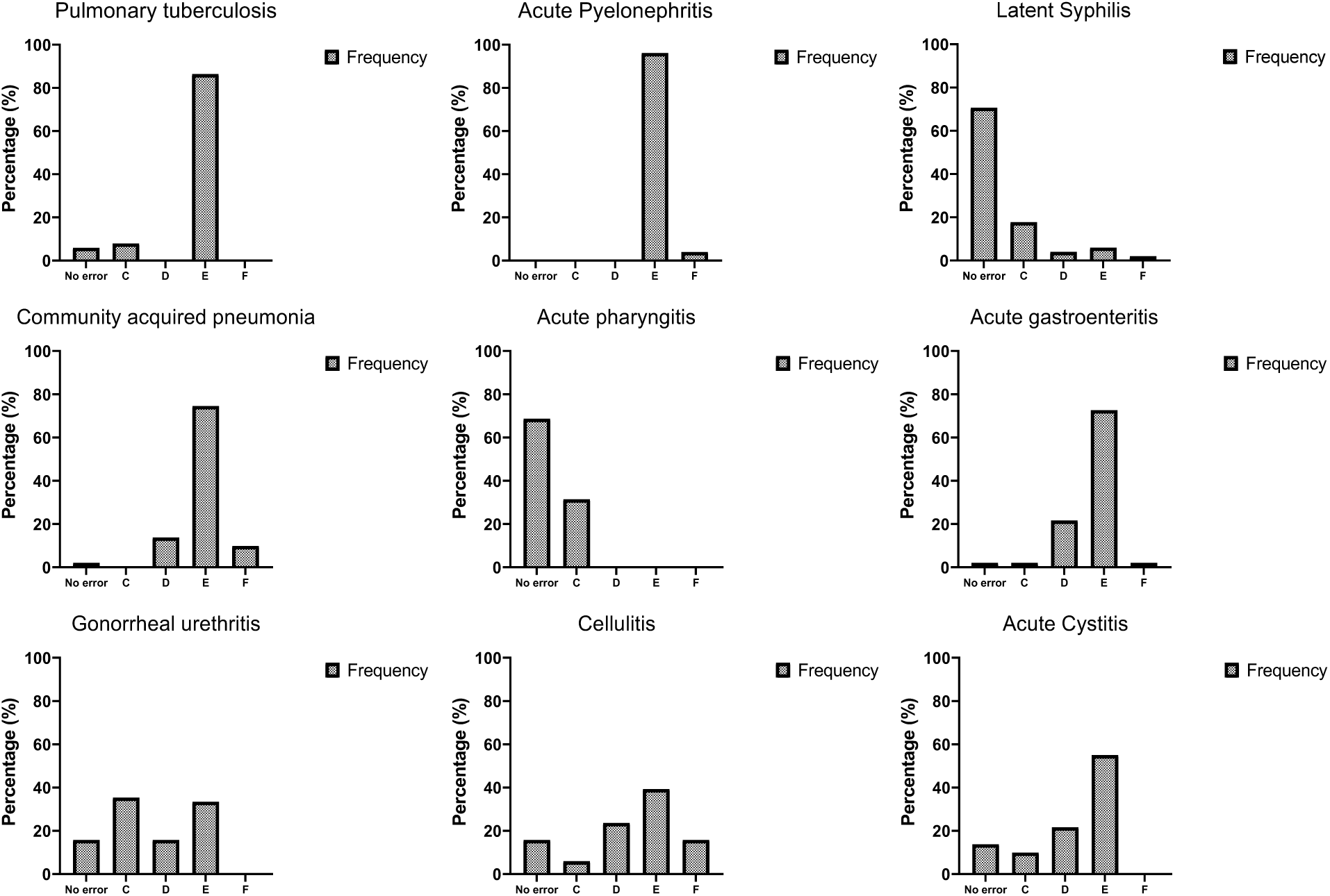
Antibiotic prescription errors severity ratio (NCC MERP Index) in each OSCE station.

### Correlation and linear regression results

The relationship between the clinical competence and antibiotic prescription errors is not quite linear (*R*^2^=0.11, Residual 0 ± 0.99) (Figure 4). Pearson correlation (r=-0.33, p<0.05, CI95% -0.57 to -0.07), and partial correlation controlling for the effect of gender and time elapsed since graduation from medical school (r=-0.39, p<0.01, CI95% -0.625 to -0.118) were computed. In addition, the correlation increases to r=-0.68 with the Spearman correction for attenuation(59). Two outliers were detected and excluded, with a 4.07 and 2.81 anomaly index.

**Figure 4.**
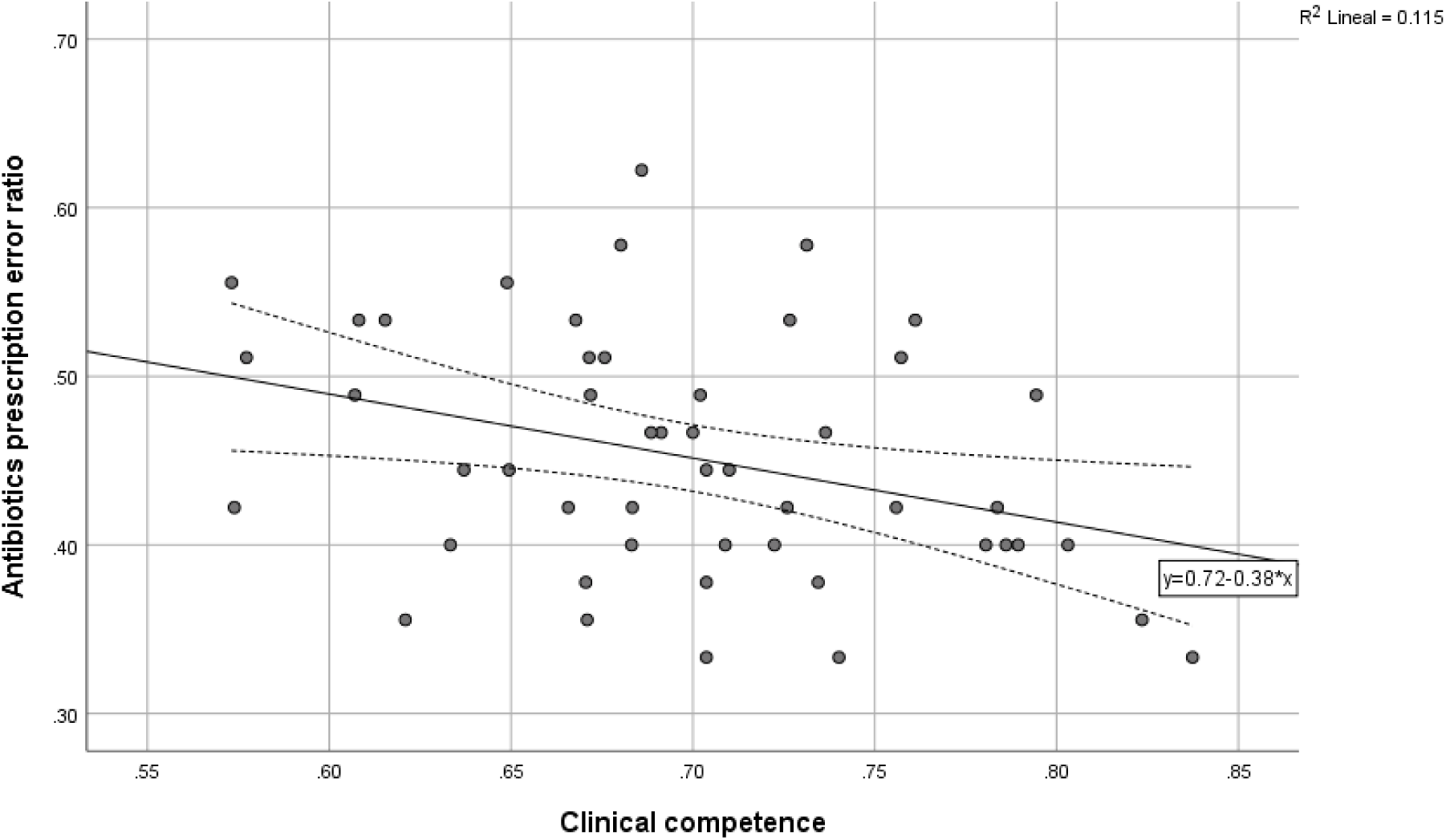
Scatter plot of antibiotic prescription errors ratio and clinical competence. The relationship between the clinical competence and the antibiotic prescription errors is linear (n=49, *β*_0_=-17.085, *β*_1_=-0.33, p<0.05, CI95%=-31.005, -3.16, *R*^2^=0.11, Residual 0 ± 0.99). The regression line was fitted with least square regression.

The clinical competence components that showed a correlation with prescription antibiotic errors ratio were the therapeutic plan (r=-0.454, p<0.05, CI95% -0.713 to -0.135) and the patient’s assessment (r=-0.351, p<0.05, CI95% -0.585 to -0.097). The prescription (r=-0. 627, p<0.05, CI95% -0.771 to -0.46) and the global assessment of knowledge and skills (r=-0.45, p<0.05, CI95% -0.68 to -0.192) items also showed significant correlations with prescription antibiotic errors ratio.

To know if the clinical competence was useful to predict the antibiotic prescription errors, we used a linear regression (*β*_0_=-17.085, *β*_1_=-0.33, p<0.05, CI95%=-31.005, -3.16, *R*^2^*=* 0.11).

### Exploratory factorial analysis

An exploratory factorial analysis (EFA) was done to describe the factors that correlates with the antibiotic prescription errors ratio. Two factors explained 69% of variance with maximum likelihood extraction method.

Factor 1 comprises anamnesis, physical examination, communication and patient and Factor 2 comprises therapeutic plan, prescription, diagnosis, and laboratory and imaging tests. We labeled factor 1 socio-clinical skills and factor 2 diagnostic-therapeutic skills. The oblique rotation was considered, although the orthogonal rotation achieve a simple structure and easier interpretation(60).

Figure 5 shows a factor plot in rotated factor space with the equamax method. The items are organized in the common factor space and shows its contributions to each factor, global assessment of knowledge and skills is the item that contributes to the 2 factors and is a complex variable. Therapeutic plan item has a highest contribution to factor 1 and communication to factor 2.

**Figure 5.**
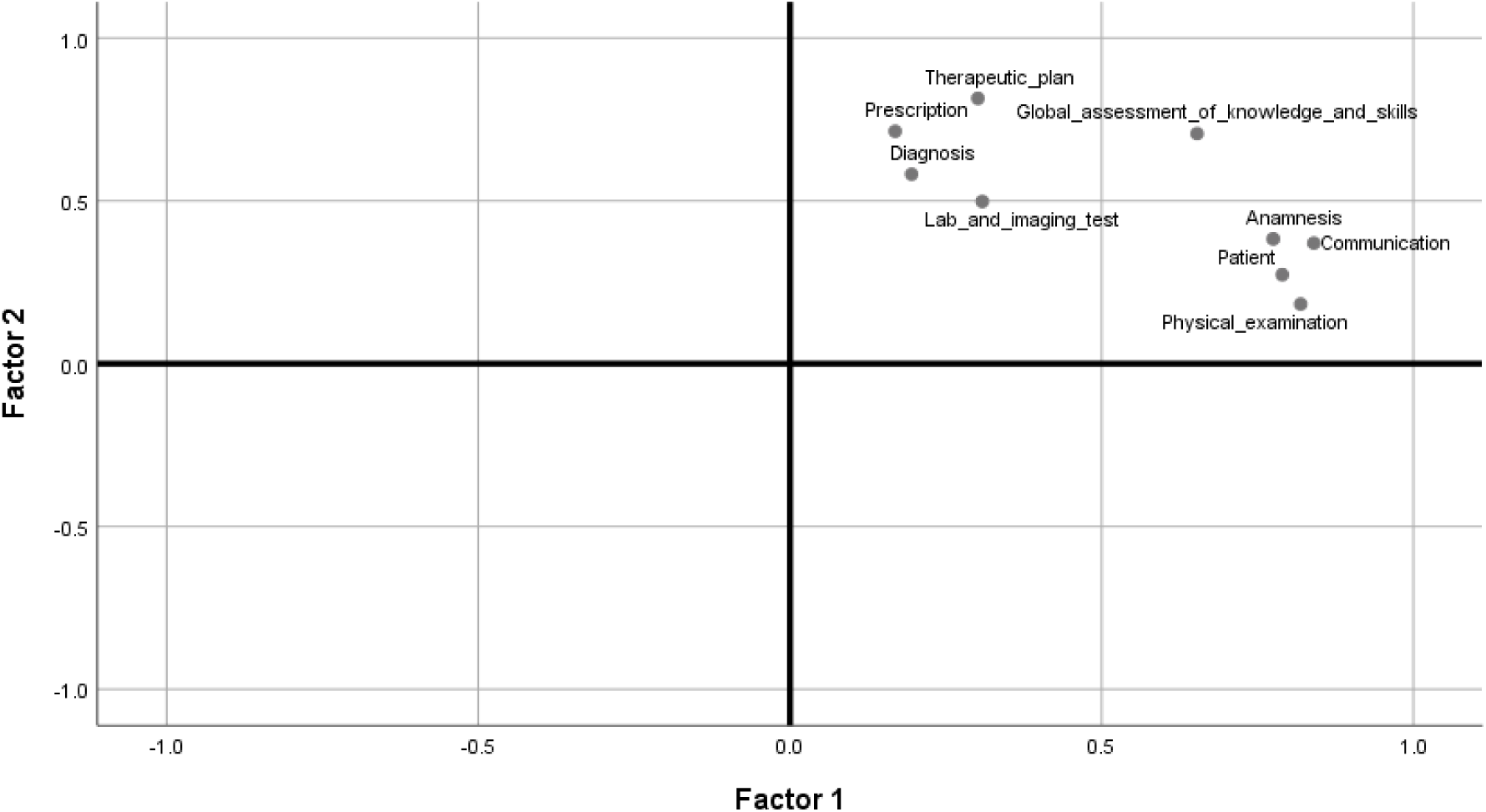
Factor plot in rotated factor space of clinical competence components and OSCE items. Factor 1 (socio-clinical skills) is a cluster by anamnesis, physical examination, communication and patient variables and Factor 2 (diagnostic-therapeutic skills) is a cluster by therapeutic plan, prescription, diagnosis and lab and imaging test variables. Varimax extraction method and equamax method.

Factor 2, diagnostic-therapeutic skills was correlated with antibiotic prescription error ratio (r=-0.536, p<0.001).

## Discussion

This study contributes to the necessity of antibiotics prescription assessment in junior physicians and identify the key learnings objectives. Our results are consistent with qualitative research in United Kingdom(61). However, we did not find a study to measures the clinical competence with an OSCE to the aim of study antibiotics prescriptions errors.

Our results show a negative correlation between the clinical competence and antibiotic prescription error ratio. The strength of association increased when we corrected the attenuation and considered sex and months since the physician’s graduation from medical school.

Our OSCE is reliable because its internal consistency measured with Cronbach’s alpha (across items 0.927 and across station 0.774) and generalizability coefficient (0.84) is higher than reported values of the systematic review of the literature (alpha across items 0.78 and alpha across station 0.66, G coefficient 0.12–0.86) (17,19)

Despite of this OSCE is aim to infectious diseases, our score is similar to a study in 7 generations of physicians, just at the end of their medical degree(57). Moreover, in Swiss and United States of America had approximate scores(62,63).

The antibiotic prescription error ratio in this study is higher than the international literature reported(3,4,11,64,65), but similar to national reported (5,66). In a prospective cohort study, the antibiotics prescription error ratio was 18% in patients hospitalized in internal medicine. Moreover, patients with adverse drug events (ADEs) had more antibiotics prescriptions errors. The most common infectious diseases were urinary tract infection and pneumonia(64). These and recommended diagnoses in Delphi’s consensus from Netherlands(67) by were included in our OSCE.

One of the objectives of the study was to find items and attributes of clinical competence that are associated with antibiotic prescription error ratio. The therapeutic plan, which is a component of clinical competence, has a negative correlation with antibiotic prescription errors. The current score of clinical competence only explains 11% of antibiotic prescription errors ratio in the linear model. Of note, most of the antibiotic prescription errors severity fell in category E (65.2%).

In addition, we introduced prescription item in synthetic guidelines for the rater in the OSCE to assess the medical prescription making process. The prescription item has a strong negative correlation with antibiotic prescription errors and high internal consistency with other items. In the future, the prescription item could be included in the components of clinical competence score to get a better assessment.

### Limitations

The medical residents voluntarily participated in this study and we did not assess medical residents who refused the test. Moreover, the institutions selected the residents previously with their own selection process and they may have higher levels of the evaluated construct than the general population of junior residents in Mexico. For these reasons, the score may not be a representative measure of the clinical competence of all general physicians.

Another potential source of bias is medical residents previous experience with OSCE. The effect on scores from medical residents with previous experience with OSCE remain unknown, although the evidence shows that it does not modify subsequent performance scores(68). The previous experience depends on the university of origin and its assessment methods.

In addition, the OSCE is a simulation and may be inflexible. The OSCE and subsequent performance had weak correlation(69). The performance in superior years of specialty course is unknown.

The medical specialty with more residents in the OSCE was pediatrics. Although the first-year medical residents had not started their courses yet; they might be biased by their vocational orientation.

The sample size is almost the absolute minimum to an EFA, and the solution should be interpreted with caution. However, the conditions to achieve good quality results fits with our values (2 factors, significant loadings of ≥0.5 and 9 variables) and sample size(70).

### Interpretation of results

Many factors influence prescription errors: lack of knowledge regarding drug therapy, lack of knowledge about patient factors that affect drug therapy, calculations and terminology (4,71). They are preventable and are ranked according to their frequency (3,72). We measured the medication errors at the prescribing stage in antibiotics because of its important implications and the high frequency of errors in this group. The OSCE measured the application of knowledge in a task, and the items regarding prescription and therapeutic plans took into consideration the common factors reported in medication errors.

Our results support an association between clinical competence and antibiotic prescription error ratio. The diagnosis item was not correlated with antibiotic prescription errors, but only the therapeutic plan. A possible explanation is that most errors occur along the therapeutic reasoning process. The exploratory factorial analysis shows a moderate correlation of diagnostic-therapeutic skills with antibiotic prescription errors, socio-clinical skills are no related.

The most frequent type of antibiotic prescription error is category E (65.2%), which means that the patients would need an intervention. A second therapeutic intervention in patients with an infectious disease has many clinical consequences, it increases the cost, leads to antibiotic resistance and increases mortality (8,73,74).

The low value of clinical competence to predict antibiotics prescription errors may be is we could not measure the complex interaction of health care culture(75). More studies that measure these variables are needed to propose a good predictor model.

### Generalizability

Our results can be applied to graduated physicians who have been accepted in a pediatrics specialty course, without fatigue. The sample had the total population of National Institute of Pediatrics.

The antibiotics prescription ratio is variable between countries but is consistent in Mexico.

### Conclusions

We found a negative correlation between clinical competence and antibiotic prescription error ratio in graduated physicians who have been accepted in a medical specialty. The therapeutic plan, which is a component of clinical competence score, and the prescription skills had a negative correlation with antibiotic prescription errors. The most frequent mistakes in antibiotic prescriptions errors would need a second intervention.

Our findings contribute to the evolving understanding of medication errors in the prescription stage of antibiotics. This study adds important evidence to improve the curricula and medical education to avoid antibiotic prescription errors, thus increasing patient safety and reducing costs, mortality and antibiotic resistance.

### Funding

This study was funded by Coordination of Educational Development and Curricular Innovation (CODEIC) of the National Autonomous University of Mexico (UNAM). We do not have conflicts of interest.

## Data Availability

The data of our study is not available yet

## Acknowledgments

All authors approved the submitted version. All the authors would like to thank the staff of the Medical Education Department, Nacional Institute of Pediatrics and “Manuel Gea González” General Hospital for their support. Joshua Martínez-Domínguez is enrolled at the PECEM program of the Faculty of Medicine at UNAM and is supported by CONACyT.

## Notes

### Competing Interest Statement

The authors have declared no competing interest.

### Clinical Trial

This study is a cross-sectional study and is not a clinical trial

### Funding Statement

This study was funded by the Coordination of Educational Development and Curricular Innovation (CODEIC) of the National Autonomous University of Mexico (UNAM).
Acknowledgments
All authors approved the submitted version. All the authors would like to thank the staff of the Medical Education Department, Nacional Institute of Pediatrics and “Manuel Gea González” General Hospital for their support. Joshua Martínez-Domínguez is enrolled at the PECEM program of the Faculty of Medicine at UNAM and is supported by CONACyT.

